# Piloting a pragmatic clinical audit tool for quality improvement in rural paediatric care in northern Sierra Leone

**DOI:** 10.1101/2023.02.01.23285336

**Authors:** Muhammed O. Afolabi, Philip Ayieko, Yusupha Njie, Dickens Kowuor, Hammed H. Adetola, Bomposseh Kamara, Abdulai Berber Jalloh, Ernest Swaray, Lazarus Odeny, Richmonda M. Pearce, Mohamed H. Samai, Gibrilla Fadlu Deen, Songor Koedoyoma, Isaac G. Sesay, David Ishola, Bailah Leigh, Deborah Watson-Jones, Brian Greenwood

## Abstract

Hospital admissions and their clinical outcomes can reflect the disease burden in a population and can be utilised as effective surveillance and impact monitoring tools. Inadequate documentation of admissions and their outcomes have contributed to the poor quality of paediatric care in many health care settings in sub-Saharan Africa. We have developed and piloted a simple tool for documentation of basic, standardised patient-level information on causes of admissions, diagnoses, treatments and outcomes in patients admitted to the paediatric ward of a district hospital in a rural community in Sierra Leone.

From 1 August 2019 to 31 July 2021, we used this tool to document the admissions, treatments and clinical outcomes of 1,663 children admitted to this hospital. The majority of the children (1015, 62%) were aged between 12-59 months, were boys (942, 57%), were wasted (516, 31%), stunted (238 14%) or underweight (537, 32%). More than half of the children lived more than 1 km distance from the hospital (876/1410, 62%). Most were admitted before 4pm (1171/1626, 72%) and during weekdays (1231/1662, 74%). The highest number of paediatric admissions occurred in November 2019 and the lowest in April 2020. Severe malaria was the leading cause of admission. More than 80% of the children were successfully treated and discharged home (1356/1663, 81.5%) while 122/1663 (7.3%) died. Children aged under-five years who were underweight, and those who presented with danger signs (e.g. signs of breathing difficulty, dehydration, head injury or severe infections) had a higher risk of death than children without these features (p<0.01; p=0.03; p=0.011 and p= 0.009, respectively).

Lack of systematic documentation of medical histories and poor record keeping of hospital admissions and outcomes can be overcome by using a simple tool. Continuous use of the tool with regular audits could improve delivery of paediatric care in resource-limited settings.

## Introduction

Sierra Leone has some of the world’s poorest health indicators. In 2020, the infant mortality rate was 80 deaths per 1,000 live births, and the overall under-five mortality rate was 108 deaths per 1,000 live births, [1] placing Sierra Leone the fourth lowest country in Africa in the ranking of these child health indicators. [2] In the same year, an estimated 27,336 under-five deaths occurred in the country, largely due to preventable causes. [3] Despite the progress recorded following the multi-faceted approaches to rebuilding the country’s fragile health system which was weakened by the 11 years of civil war and the 2014-2016 Ebola outbreak [4], improving child survival remains a major public health challenge in the country. [5]

Prompt diagnosis and early provision of appropriate health care are widely recognised as important strategies to improve child survival. [6] This needs to occur at primary and secondary health care levels. In many Low or Middle Income Countries (LMIC), there is frequently incomplete documentation of patient admissions and outcomes, [6] which can hinder appropriate planning and implementation of effective service delivery. [7] Thus, improving documentation of admissions and outcomes in secondary health care facilities can support impact evaluation, exploration of links between health system inputs and outcomes and provide critical information on geographical variations in quality and outcomes of care. [8, 9] In addition, reliable hospital data can provide valuable insights on the disease burden in a population and can be utilised as effective surveillance and impact monitoring tools. The data provided by these records can serve as an entry point to the investigation of system weaknesses and failures, assessment of the quality of health systems and can be used as a prism through which determinants of health system performance can be interrogated. [8]

The scarcity of reliable data on births, deaths and causes of death in many LMIC has led to heavy reliance on extrapolating data obtained from a few populations that are under demographic surveillance and from verbal autopsies. [8] More effective use of quality hospital data has been advocated by global health organisations in order to understand disease burden in a specific community, enabling prioritisation of resource allocations [10] and facilitating achievement of universal health coverage.

Towards addressing this need, we have developed and piloted a simple tool for documentation of basic, standardised patient-level information on causes of admissions, diagnoses, treatments and outcomes in a paediatric ward of a district hospital in a rural community in Sierra Leone. In this paper, we describe the pattern of admissions and clinical outcomes of the children admitted to the paediatric ward of the Kambia Government Hospital, Sierra Leone, using this tool continuously over a two-year period. During this study, the COVID-19 outbreak spread across Sierra Leone and lockdowns and travel restrictions were imposed by the government as public health measures.

Therefore, we also assessed the impact of the COVID-19 outbreak on the paediatric admissions and outcomes in the hospital.

## Materials and Methods

### Study population

Kambia is located in the north-west region of Sierra Leone, about 200 km from Freetown, the capital city. Kambia is one of the communities that was heavily affected by the country’s civil war (1991-2002) and the West African Ebola outbreak in 2014-2016. [11] Kambia district has one of the highest childhood morbidities and mortalities among the 14 administrative districts of Sierra Leone. [12] Top amongst the causes of death and illness in children are malaria, pneumonia and diarrheal diseases. Government primary health care services consist of peripheral health units, which are run by nurses and community health officers. Remote and rural settlements are served informally by unregulated mobile drug vendors and traditional practitioners. [13]

Kambia Government Hospital (KGH) is the only public health centre providing primary and secondary health care for a population of more than 60,000 under-five children. [14] The hospital has four wards (adult male, adult female, paediatric and maternity), one surgical theatre and two outpatient clinic rooms. The paediatric ward is run primarily by nurses and community health officers. However, at the time of this study, a paediatrician and a paediatric nurse were employed to provide specialist care for the paediatric ward by the EBOVAC-Salone project which was evaluating a prophylactic Ebola vaccine regimen (clinicaltrials.gov NCT02509494) in Kambia district. [15, 16] Prior to the deployment of the tool described in this paper, few records were kept for patients accessing care at the hospital. Clinical histories and care plans were usually documented in ad-hoc note books procured by parents/caregivers on arrival at the hospital ward with their sick children. Consequently, paediatric admissions and outcomes were often incompletely documented in the hospital register. To improve collection of individual patient-level data on hospital admissions, we developed a simplified, user-friendly, paper-based tool which contained sections covering basic socio-demographic information of a patient, including age, gender, home address, source of referral, date and time of admission, chief medical complaints, findings from clinical examinations and tests conducted at admission, results from subsequent clinical and laboratory investigations, working/definitive diagnoses, management and treatment outcomes. The tool (Supplementary file S1) was pre-tested for validity among health workers who were not involved in the study and revised to incorporate feedback received from the pre-test. Practical training and demonstrations on how to use the tool to collect information from parents/care-givers were provided for the hospital staff. Hospital staff members including nurses and community health workers were also trained on basic medical record keeping and were helped to set up an analogue medical record unit in the paediatric ward of the hospital. Oversight was provided by the attending physicians (MOA and HHA) to ensure that the tool was used to collect complete and legible data in a timely fashion.

### Ethical considerations

Ethical approvals for the study were obtained from the Research Ethics Committee of the London School of Hygiene & Tropical Medicine and the Sierra Leone Ethics and Scientific Review Committee. Written permission for the study was obtained from the Management of the Kambia Government Hospital, an arm of the Sierra Leone Ministry of Health and Sanitation. Written informed consent was obtained from the parents/care-givers before the tool was used to collect their child’s health information. In addition, assent was obtained from children aged seven years and above. Personal data collected about the hospitalised children were anonymised, kept confidential and held in compliance with international data privacy protection laws and regulations.

### Statistical analysis

Hospital data collected during the implementation period were double-entered in an OpenClinica database at the Kambia COMAHS/LSHTM research centre, cleaned and then analysed using descriptive and inferential statistics. All statistical analyses were conducted using Stata 16 Statistical Software (Stata Corp, USA). Information on the clinical indices (presentations, investigations, interventions, complications, and outcomes) and service indices (such as the duration of admission) were represented through graphical charts. We used Chi squared tests to evaluate the associations between individual characteristics of the hospitalised children during the study and categorised the clinical outcomes into four levels – discharged alive, referred, discharged against medical advice by parents and death during hospitalisation. Unadjusted logistic regression was performed to examine risk factors for death among paediatric admissions by assessing the association between death and each risk factor in turn. Next, we conducted multivariable regression analysis for death adjusted for time of the day during which admission occurred, presence of danger signs (such as signs of breathing difficulty, dehydration, head injury and severe infections) during admission and weight-for-age and weight-for-height Z scores which had shown significant associations with mortality in the unadjusted logistic regression, based on two-tailed p value cut-off < 0.05.

## Results

We administered the tool from 1 August 2019 to 31 July 2021 to a total of 1663 children out of 2068 children admitted into the ward during the same period, representing an 80% response rate. The main reason for failure to administer the tool to all admitted children during this period was an early discharge from the hospital before consent could be obtained from the parents/caregivers.

### Characteristics of hospitalised children

Table 1 summarises the socio-demographic characteristics of the 1663 children enrolled into the study. Overall, 1015 (62%) were aged 12-59 months and 942 (57%) were boys. Less than 10% of the children (112, 9%) were severely malnourished based on the mid-upper arm circumference (MUAC) measurements of <11.5cm, while 516 (31%) were wasted, 238 (14%) were stunted and 537 (32%) were underweight. Over half of the children lived more than 1 kilometre from the hospital (876/1410, 62%), and most were admitted before 4pm (1171/1626; 72%) and during weekdays (1231/1662; 74%).

**Table 1:** Socio-demographic characteristics of study participants, KGH, 2019-2021.

|  | **N = 1663**<br>**n (%)** |
| --- | --- |
| Age groups |  |
| 0-27 days (neonates) | 141(9) |
| 1-<12 months (infants) | 312(19) |
| 12-59 months | 1015(62) |
| 5-14 years | 181(11) |
| Sex |  |
| Male | 942(57) |
| Female | 720(43) |
| MUAC |  |
| Below 11.5cm | 112(9) |
| Above 11.5cm | 1148(91) |
| WFA |  |
| < -2 SD | 516(31) |
| >-2 SD | 1006(60) |
| Missing | 141(8) |
| HFA |  |
| < -2 SD | 238(14) |
| >-2 SD | 1144(69) |
| Missing | 281(17) |
| WFH |  |
| < -2 SD | 537(32) |
| >-2 SD | 510(31) |
| Missing | 616(37) |
| *Proximity of residence to KGH |  |
| Within 1 km | 534(38) |
| More than 1 km | 876(62) |
| †Time of admission |  |
| Before 4 pm | 1171(72) |
| After 4 pm | 455(28) |
| Admission day |  |
| During weekdays | 1231(74) |
| During weekends | 431(26) |
| Sources of referral |  |
| Within KGH | 114(7) |
| Self/parent referred | 1069(64) |
| Other facility | 478(29) |

WFA = Weight-for-age; HFA= Height-for-age; WFH = Weight-for-height
\*Number of children whose home addresses were correctly provided by parents/caregivers = 1410
†Number of children whose times of admission were documented = 1626

### Seasonality of admission

Over the study period, the highest number of admissions was recorded in November 2019 and lowest admissions occurred in April 2020 (Figure 1). These patterns of admissions mirror the changes in the weather conditions in the study area where the dry season typically runs from November to March and the rainy season from April to October with a peak of rainfall in August. However, the seasonal pattern of admissions may have been modified during the audit in 2020 by the COVID-19 pandemic, with the first imported case of COVID-19 reported on 30 March 2020.

**Figure 1:**
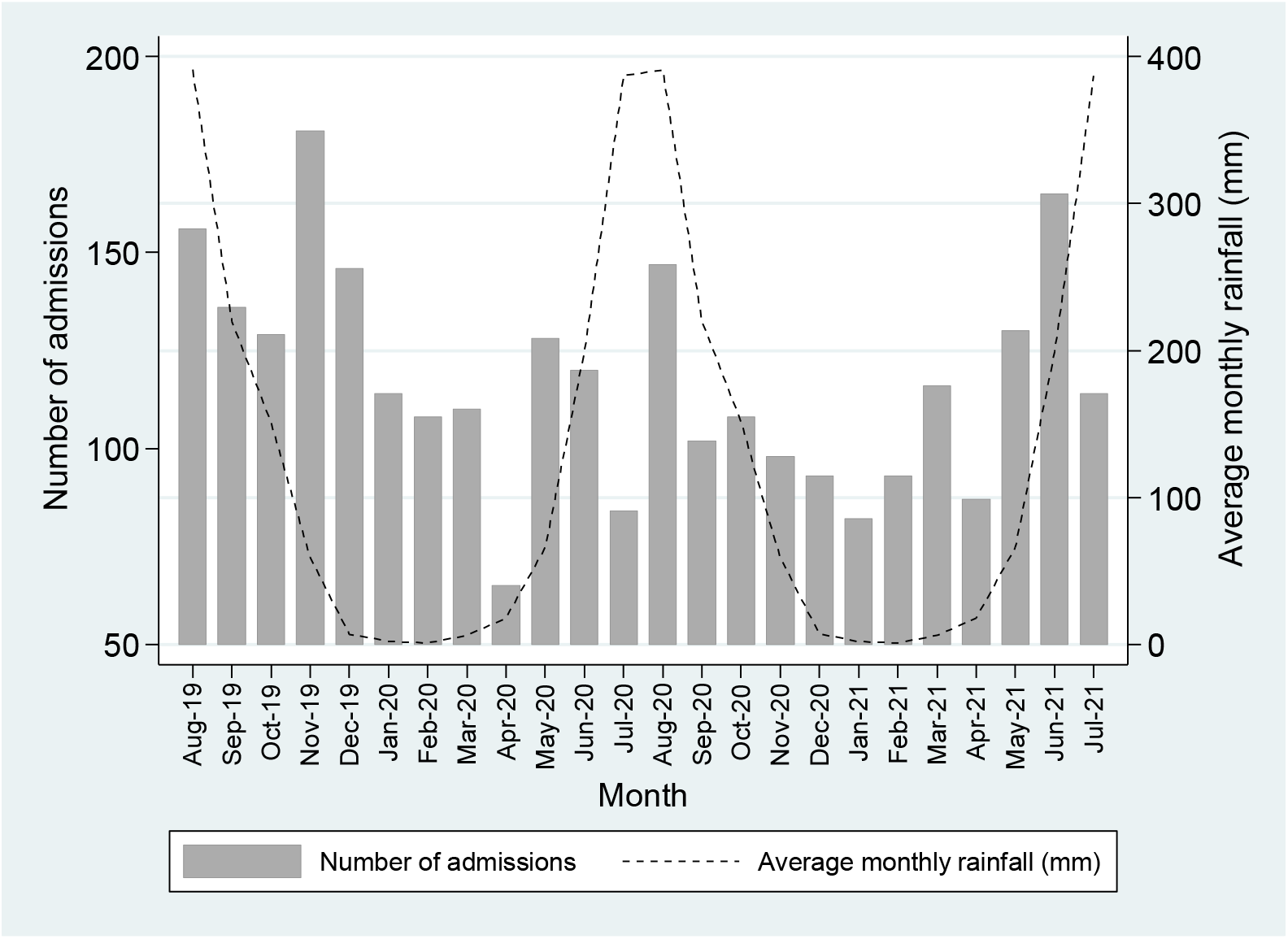
Seasonal pattern of admissions to the KGH paediatric ward, Aug 2019-Jul 2021

### Impact of COVID-19 lockdown

The national lockdowns and restrictions of movements imposed in Sierra Leone in the wake of COVID-19 outbreak had an impact on paediatric admissions. There were two 3-day national lockdowns between 5–7 April and 3–5 May 2020. From 14 April to 4 July, there were movement restrictions within districts across the country. From 22 March to 22 July 2020, international commercial air travel was suspended. [17] During these periods, schools were closed and considerable reductions in paediatric admissions were observed during and around the periods of these lockdowns, with the nadir recorded two weeks after the first national lockdown (Figure 2).

**Figure 2:**
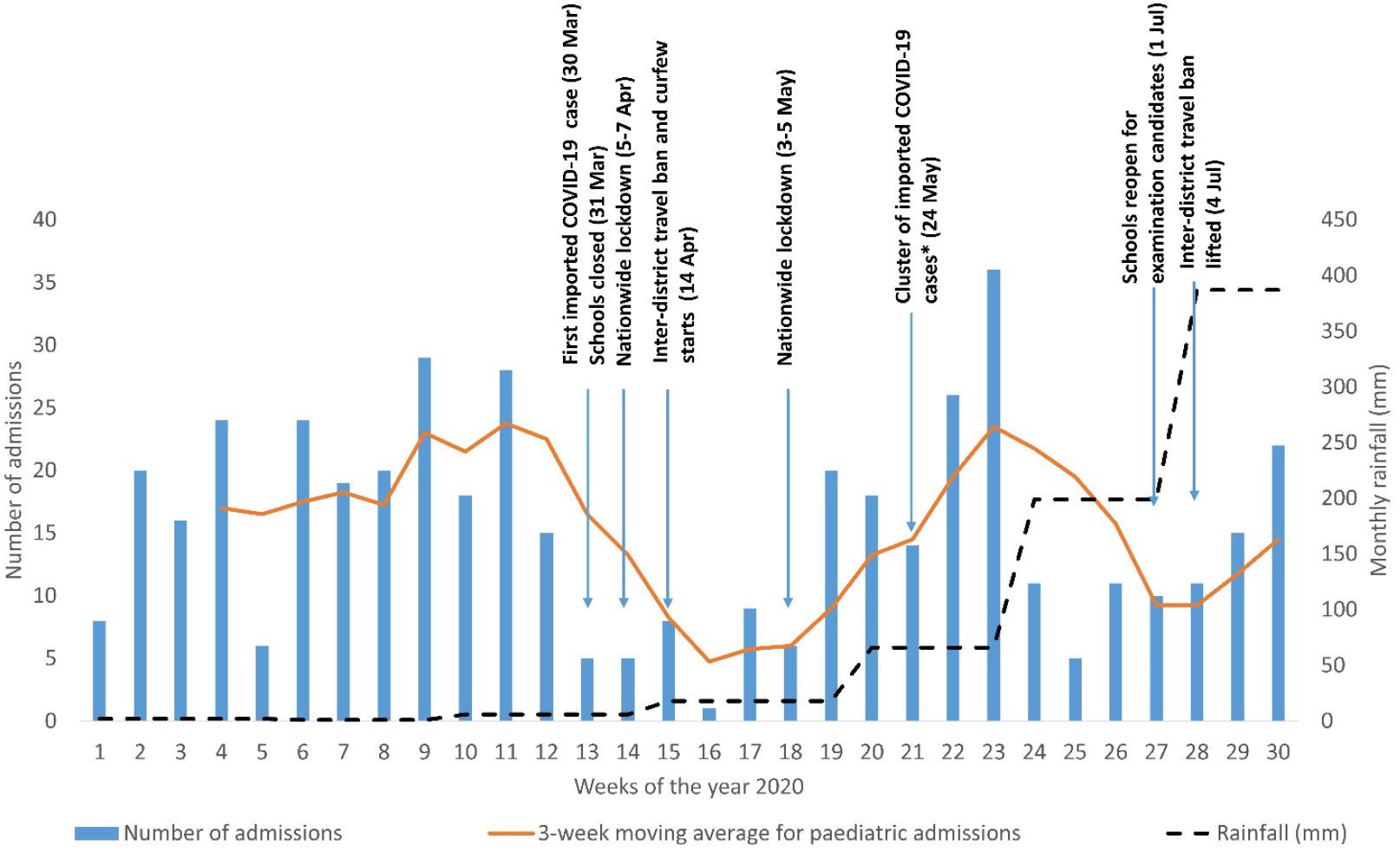
Impact of COVID-19 pandemic on admissions among study participants, KGH, 2019-2021

**Figure 3:** Clinical diagnoses among study participants, KGH, 2019-2021

### Admission diagnosis

Out of the 1663 paediatric admissions, 1,053 (63.3%) had a single admission diagnosis; 474 (28.5%) had two co-morbid diagnoses, and 102 (6.1%) had three or more diagnoses. There was no documentation of a diagnosis in the admission records of 34 (2%) participants. After assigning a primary diagnosis for each of the 576 participants with co-morbid illnesses, the most common diagnoses among all participants were malaria 1089 (65%); asphyxia 86 (5%); malnutrition 55 (3%); sepsis 51 (3%); and surgical conditions 51 (3%). The leading clinical phenotypes for malaria were severe malarial anaemia 498 (30% of all admission), severe malaria not meeting other criteria: 383 (23%) and cerebral malaria 106 (6%) [Table 2].

**Table 2:** Primary diagnoses among children admitted to KGH, 2019-2021.

|  | Age in years |  |  |  | Total<br>n (%) |
| --- | --- | --- | --- | --- | --- |
|  | < 1 year<br>n (%) | 1-4 years<br>n (%) | 5-14 years<br>n (%) | No age<br>documented<br>n (%) |  |
| **Primary admission**<br>**diagnosis (N = 1663)** |  |  |  |  |  |
| Severe malarial anaemia | 71 (16) | 373 (37) | 52 (29) | 2 (14) | 498 (30) |
| Severe malaria | 58 (13) | 267 (26) | 54 (30) | 4 (29) | 383 (23) |
| Cerebral malaria | 12 (3) | 73 (7) | 21 (12) | - | 106 (6) |
| Severe malaria with<br>respiratory distress | - | 9 (1) | 1 (1) | - | 10 (1) |
| Non-severe malaria | 20 (4) | 63 (6) | 8 (4) | 1 (7) | 92 (6) |
| Asphyxia | 83 (18) | 1 (0) | - | 2 (14) | 86 (5) |
| Malnutrition | 15 (3) | 40 (4) | - | - | 55 (3) |
| Sepsis | 33 (7) | 14 (1) | 3 (2) | 1 (7) | 51 (3) |
| Surgical conditions/ burns/<br>trauma/ accidents | 6 (1) | 25 (2) | 19 (10) | 1 (7) | 51 (3) |
| Severe pneumonia | 29 (6) | 12 (1) | - | 1 (7) | 42 (3) |
| Anaemia | 11 (2) | 27 (3) | 3 (2) | - | 41 (2) |
| Acute gastroenteritis | 16 (4) | 22 (2) | 1 (1) | - | 39 (2) |
| Febrile convulsion | 4 (1) | 18 (2) | 2 (1) | - | 24 (1) |
| Neonatal sepsis | 19 (4) | - | - | 1 (7) | 20 (1) |
| Prematurity | 12 (3) | - | - | - | 12 (1) |
| Non-severe pneumonia | 9 (2) | 2 (0) | - | - | 11 (1) |
| HIV | 1 (0) | 6 (1) | - | - | 7 (0) |
| Other infectious diagnoses | 18 (4) | 11 (1) | 8 (4) | - | 37 (2) |
| Other non-infectious<br>diagnoses | 13 (3) | 18 (2) | 4 (2) | - | 35 (2) |
| > 1 diagnosis but primary<br>diagnosis not determined | 14 (3) | 11 (1) | 3 (2) | 1 (7) | 29 (2) |
| Diagnosis not documented | 9 (2) | 23 (2) | 2 (1) | - | 34 (2) |
| **Total** | **453(100)** | **1015(100)** | **181(100)** | **14(100)** | **1663(100)** |

Malaria diagnosis using a rapid diagnostic test (RDT) and microscopy was performed on most children admitted to the ward in whom a positive test was obtained in 72.3% (1193/1649). When disaggregated by age group, malaria positivity by RDT and microscopy was highest among children aged 1-4 years (853/1015, 84%), followed by children aged 5-14 years (147/181, 81.2%). Of the 1193 children diagnosed with malaria by RDT and microscopy, 79.8% (952/1193) had severe malaria. Of the 952 children with severe malaria, 553/952 (58.1%) had severe anaemia; prostration was observed in 539/952 (56.6%), cerebral malaria in 147/952 (15.4%), and respiratory distress in 1.6% (15/952) [Table 3]. When disaggregated by age group, prostration, impaired consciousness, multiple convulsions and cerebral malaria were observed more frequently among 1-4 years old children than among other age groups (p<0.001). Although respiratory distress was disproportionately higher among 1-4 years old children than other age groups, this did not reach statistical significance (p=0.192) [Table 4].

**Table 3:** Malaria diagnosis and severe malaria clinical syndromes according to the age of paediatric admissions at KGH, 2019-2021

|  | Age |  |  |  |
| --- | --- | --- | --- | --- |
|  | < 1 year | 1-4 years | 5-14 years | Total |
|  | n (%) | n (%) | n (%) | n (%) |
| **Number of hospital admissions** | **N = 453** | **N = 1015** | **N = 181** | **N = 1649\*** |
| **RDT/ microscopy confirmed malaria** |  |  |  |  |
| Positive | 193(42.6) | 853(84) | 147(81.2) | 1193 (72.3) |
| Negative | 87(19.2) | 109(10.7) | 13(7.2) | 209 (12.7) |
| Malaria test not done | 173(38.2) | 53(5.2) | 21(11.6) | 247 (15.0) |
| **Malaria severity** | **N= 193** | **N = 853** | **N = 147** | **N = 1193\*** |
| Uncomplicated malaria | 55(28.5) | 162(19.0) | 24(16.3) | 241 (20.0) |
| Severe malaria† | 138(71.5) | 691(81.0) | 123(83.7) | 952 (79.8) |
| **Clinical features of severe malaria** | **N =138** | **N = 691** | **N = 123** | **N = 952\*** |
| Severe anaemia‡ | 77(55.8) | 415(60.0) | 61(49.6) | 553 (58.1) |
| Prostration | 76(55.1) | 390(56.4) | 73(59.3) | 539 (56.6) |
| Impaired consciousness | 45(32.6) | 302(43.7) | 55(44.7) | 402 (42.2) |
| Multiple convulsions | 16(11.6) | 69(10.0) | 15(12.2) | 100 (10.5) |
| Cerebral malaria | 16(11.6) | 104(15.0) | 27(22.0) | 147 (15.4) |
| Respiratory distress | 1(0.7) | 12(1.72) | 2(1.7) | 15 (1.6) |

\*14 children who did not have documentation of age were not included in analysis for age distribution of malaria diagnosis and for clinical features by age.
†Severe malaria defined as acute malaria with signs of organ dysfunction and/or high level of parasitaemia characterised by impaired consciousness, hypoglycaemia, shock, pulmonary oedema, etc
‡ Severe anaemia is defined by haemoglobin concentration of $< 5$ g/dl or a haematocrit of $< 15\%$ .

**Table 4.**
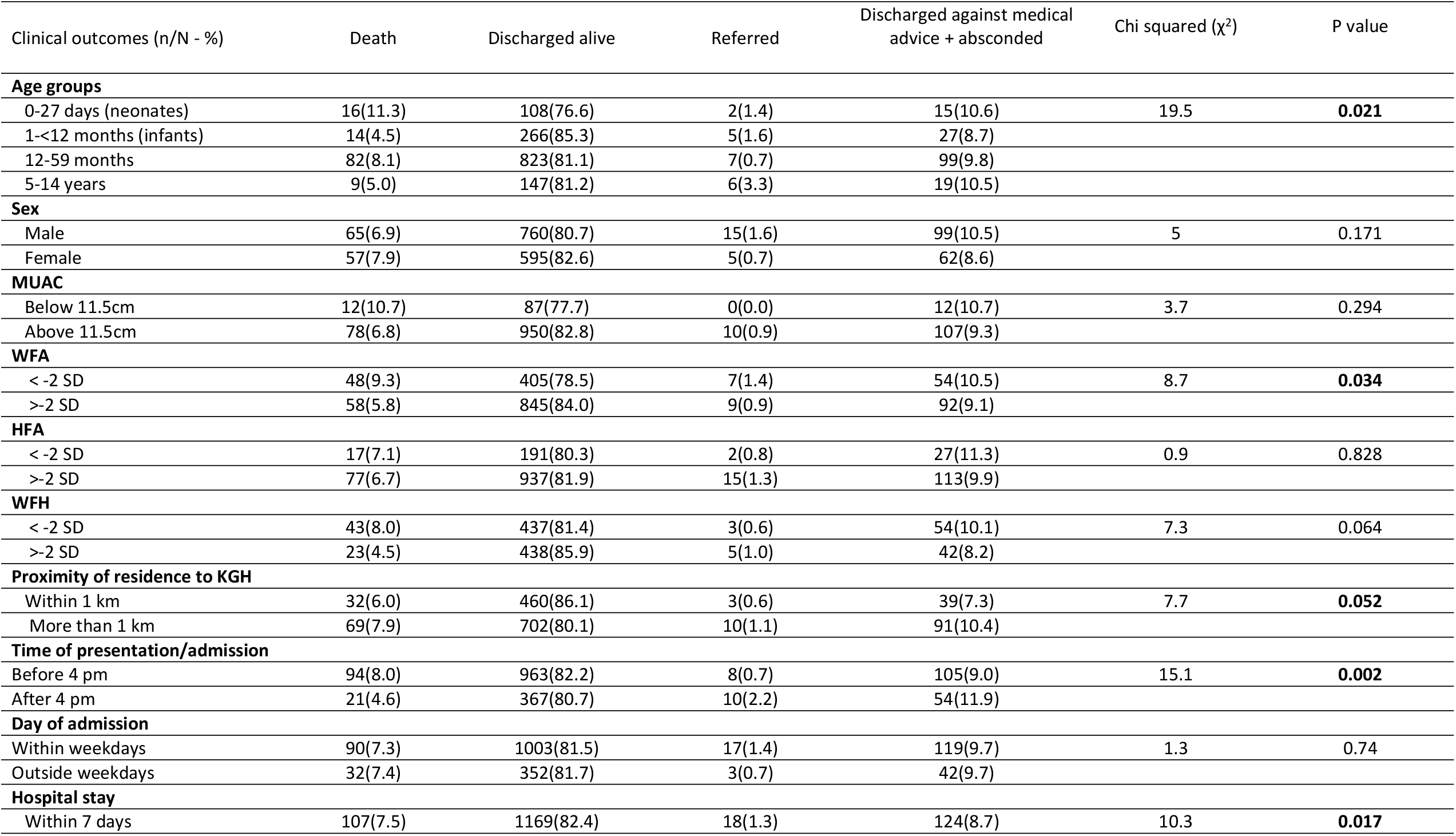
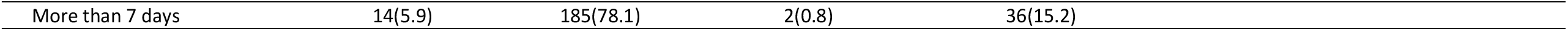
Clinical outcomes of children admitted to the paediatric ward, KGH, 2019-2021.

| Clinical outcomes (n/N - %) | Death | Discharged alive | Referred | Discharged against medical advice + absconded | Chi squared ($\chi^2$) | P value |
| --- | --- | --- | --- | --- | --- | --- |
| Age groups |  |  |  |  |  |  |
| 0-27 days (neonates) | 16(11.3) | 108(76.6) | 2(1.4) | 15(10.6) | 19.5 | 0.021 |
| 1-<12 months (infants) | 14(4.5) | 266(85.3) | 5(1.6) | 27(8.7) |  |  |
| 12-59 months | 82(8.1) | 823(81.1) | 7(0.7) | 99(9.8) |  |  |
| 5-14 years | 9(5.0) | 147(81.2) | 6(3.3) | 19(10.5) |  |  |
| Sex |  |  |  |  |  |  |
| Male | 65(6.9) | 760(80.7) | 15(1.6) | 99(10.5) | 5 | 0.171 |
| Female | 57(7.9) | 595(82.6) | 5(0.7) | 62(8.6) |  |  |
| MUAC |  |  |  |  |  |  |
| Below 11.5cm | 12(10.7) | 87(77.7) | 0(0.0) | 12(10.7) | 3.7 | 0.294 |
| Above 11.5cm | 78(6.8) | 950(82.8) | 10(0.9) | 107(9.3) |  |  |
| WFA |  |  |  |  |  |  |
| < -2 SD | 48(9.3) | 405(78.5) | 7(1.4) | 54(10.5) | 8.7 | 0.034 |
| >-2 SD | 58(5.8) | 845(84.0) | 9(0.9) | 92(9.1) |  |  |
| HFA |  |  |  |  |  |  |
| < -2 SD | 17(7.1) | 191(80.3) | 2(0.8) | 27(11.3) | 0.9 | 0.828 |
| >-2 SD | 77(6.7) | 937(81.9) | 15(1.3) | 113(9.9) |  |  |
| WFH |  |  |  |  |  |  |
| < -2 SD | 43(8.0) | 437(81.4) | 3(0.6) | 54(10.1) | 7.3 | 0.064 |
| >-2 SD | 23(4.5) | 438(85.9) | 5(1.0) | 42(8.2) |  |  |
| Proximity of residence to KGH |  |  |  |  |  |  |
| Within 1 km | 32(6.0) | 460(86.1) | 3(0.6) | 39(7.3) | 7.7 | 0.052 |
| More than 1 km | 69(7.9) | 702(80.1) | 10(1.1) | 91(10.4) |  |  |
| Time of presentation/admission |  |  |  |  |  |  |
| Before 4 pm | 94(8.0) | 963(82.2) | 8(0.7) | 105(9.0) | 15.1 | 0.002 |
| After 4 pm | 21(4.6) | 367(80.7) | 10(2.2) | 54(11.9) |  |  |
| Day of admission |  |  |  |  |  |  |
| Within weekdays | 90(7.3) | 1003(81.5) | 17(1.4) | 119(9.7) | 1.3 | 0.74 |
| Outside weekdays | 32(7.4) | 352(81.7) | 3(0.7) | 42(9.7) |  |  |
| Hospital stay |  |  |  |  |  |  |
| Within 7 days | 107(7.5) | 1169(82.4) | 18(1.3) | 124(8.7) | 10.3 | 0.017 |

|  |  |  |  |  |
| --- | --- | --- | --- | --- |
| More than 7 days | 14(5.9) | 185(78.1) | 2(0.8) | 36(15.2) |

### Clinical outcomes

More than 80% of the children were treated successfully and discharged home (1356/1663, 81.5%) while 122 (7.3%) died, 93 (5.6%) absconded, and 68 (4.1%) were discharged against medical advice. Reasons for discharge against medical advice included an inability to pay the hospital bills and some children were discharged against medical advice when their parents or guardians perceived that their child was not making satisfactory clinical improvement despite the medical management instituted. Twenty children (1.2%) were referred for further management to the paediatric specialist hospital in Freetown. Referral to the specialist hospital was mainly for the management of surgical conditions such as hernia, severe burns and head injuries following a road traffic accident. Factors that influenced the clinical outcomes among the children admitted to the Kambia hospital included age group (p=0.021), weight-for-age (p=0.034), proximity of residence to the hospital (p=0.052), time of presentation or admission at the hospital (p=0.002) and duration of hospital stay (p=0.017) [Table 4].

### Risk of death

Children aged under-five years (p<0.01), those underweight (p=0.03), or who presented with danger signs (p=0.009) had a higher risk of death than children without these features (Table 4). The risk of death was greatest within 48 hours of admission and gradually tapered off after this time point (Figure 4).

**Figure 4:**
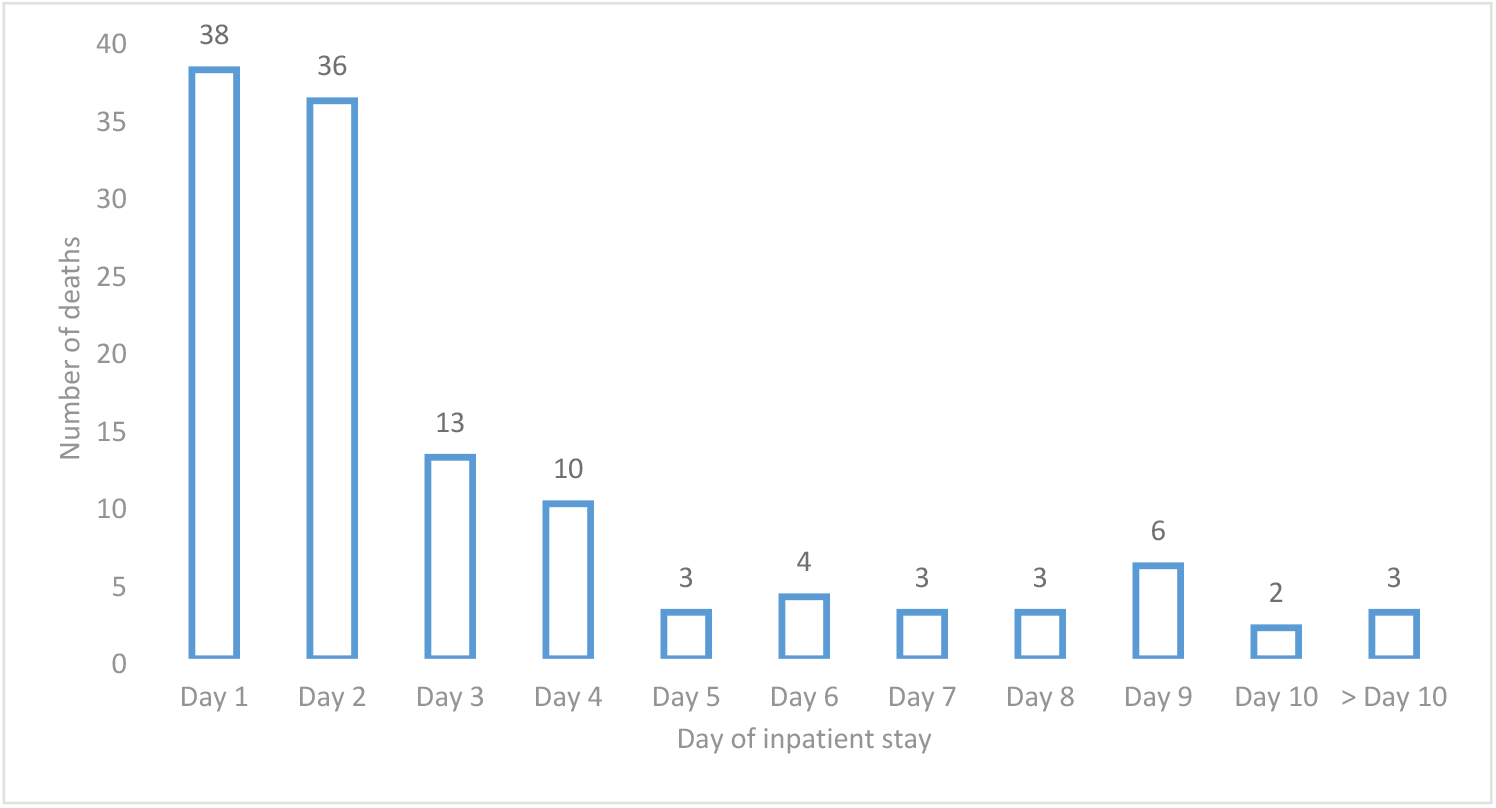
Time-to-death for admissions during the first 10 days of inpatient stay, KGH, 2019-2021

**Table 4:**
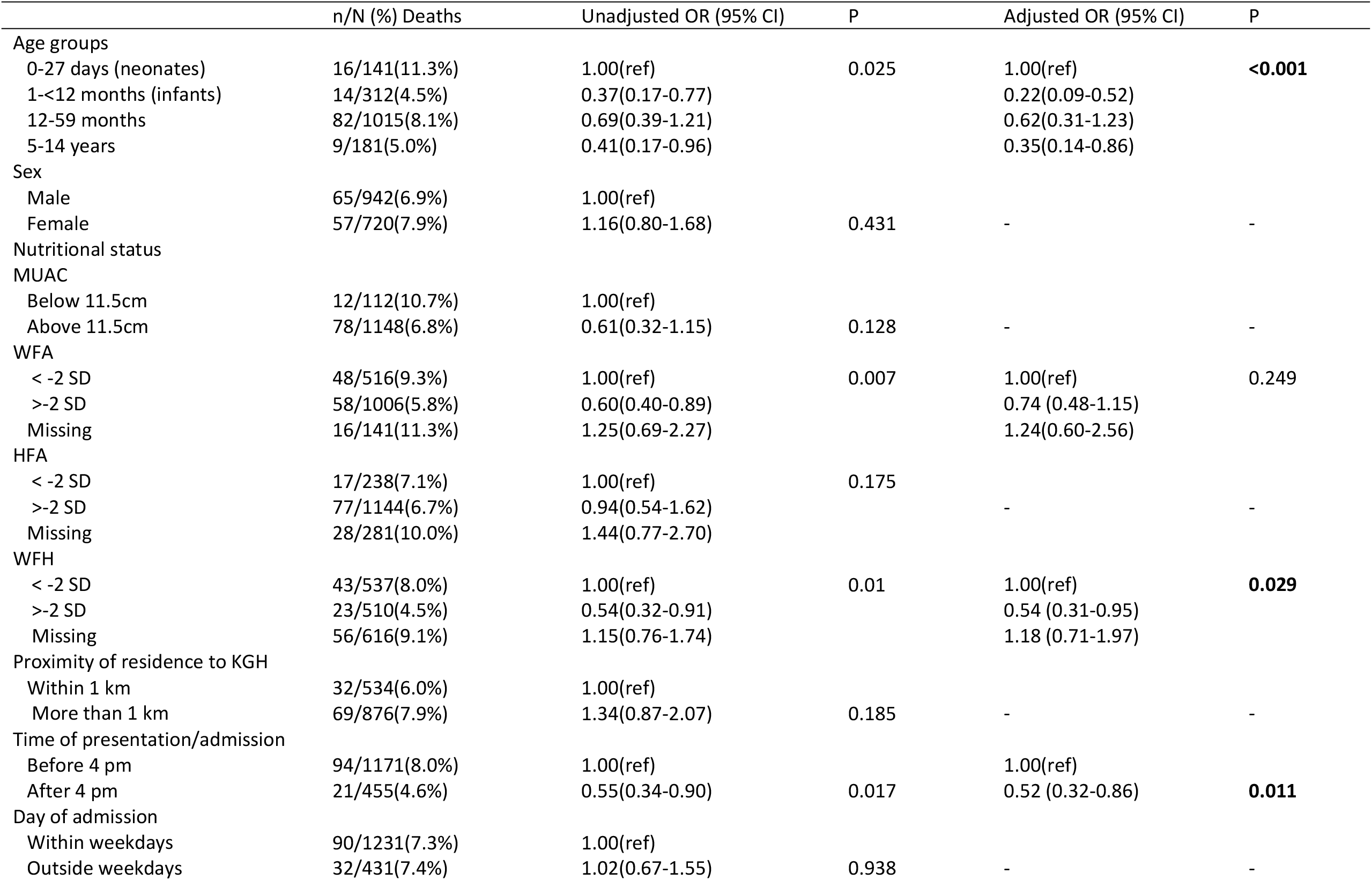
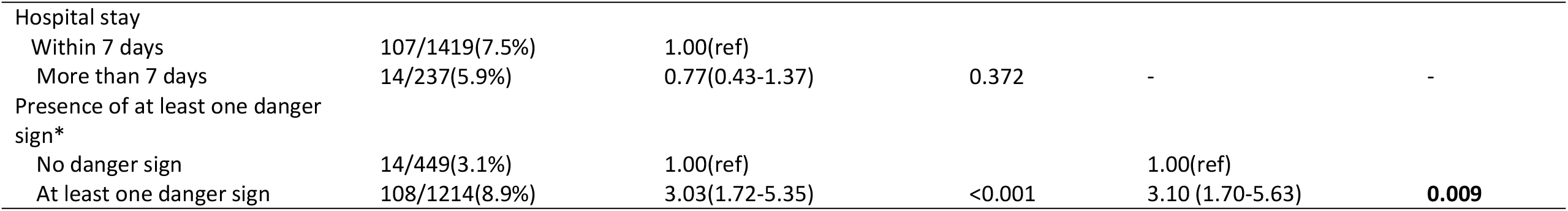
Risk factors for deaths among paediatric inpatients in KGH, 2019-2021.

## Discussion

We developed and implemented a simple tool to improve the documentation of paediatric admissions and clinical outcomes in a resource-constrained, rural hospital in northern Sierra Leone. Under-five male children constituted the majority of the children admitted to the hospital during the implementation of the tool. These demographic features are consistent with findings obtained from studies conducted in similar settings. [18, 19] Higher admission rates of boys to the hospitals in these studies were attributed to an increased vulnerability of male children to some illnesses and the prevalent African custom of placing a higher premium on care of a male child compared with a female child, because of the relatively higher social importance attached to a male child. [20] Nevertheless, the gender disparity in the admission rate was not identified as a risk factor for death in our study population. This finding differs from previous studies conducted in similar settings where mortality in some studies was higher in females [18, 21, 22] and higher in males in other studies. [19, 23]

Although, severe acute malnutrition (SAM) was not a major, primary cause of admission among the hospitalised children in our study, about a third of the children had wasting and were underweight. This nutritional status is typical of the features commonly seen in similar African settings where low rates of exclusive breastfeeding and incorrect complementary feeding practices are prevalent. [24] The difference observed in the proportions of children diagnosed with SAM based on the anthropometric findings in the population of hospitalised children could be explained by the different methods used in arriving at the diagnosis. Whilst middle-upper arm circumference (MUAC) is the anthropometric measurement recommended for the diagnosis of SAM in under-five children, [25] the use of WFH z-scores could pose diagnostic challenges to hospital staff in rural settings and might lead to variations in the proportion of children identified with SAM.

A majority of the children in our study lived more than 1 km from the hospital, although this did not translate to late presentation to the hospital. This might be because commercial motorcycles taxis, the most common means of transport in the study community, were readily available across the community and could be used to take a sick child to the hospital. More than 70% of the children presented at the hospital before 4pm and during weekdays. These findings are consistent with the results of a Gambian study. [26] Admission into the paediatric ward followed the seasonal pattern that has been widely reported in many African settings [27-32]. This is mainly because malaria still remains the leading cause of hospitalisations in these endemic countries, and given that malaria transmission is driven by environment factors such as rainfall, it is not surprising that a rise in admissions was observed during the rainy seasons. [33]

Owing to the rural location of the study hospital, which is not connected to the national electricity grid and depends largely on electricity supply from generators to power essential life-saving equipment, a point-of-care test was the major laboratory investigation deployed to confirm clinical suspicions of malaria. Similarly, because of the limited diagnostic facilities, the diagnosis of common childhood diseases such as pneumonia and meningitis depended on clinical symptoms and signs.

This practice is consistent with the WHO guidelines recommendations in the integrated management of childhood illnesses that do not focus on a single diagnosis, but on selected signs and symptoms to guide rational treatment. [34] These guidelines have improved the case-management skills of health workers in settings such as Kambia. [35]

As expected, malaria was the leading cause of paediatric admissions during the course of the tool implementation. Manifestations of severe malaria and its complications including cerebral malaria, prostration, severe malaria anaemia, and complex febrile convulsions were consistent with findings of similar studies conducted in malaria endemic countries in sub-Saharan Africa [18, 23, 28-30]; and lend credence to a WHO report that progress in malaria control has stalled despite the considerable achievements recorded in the last two decades. [36]

More than 80% of the children enrolled in this study were treated successfully and discharged home. This is a direct reflection of the improvement in the care provided by the hospital staff who were trained in Emergency Triage, Assessment and Treatment plus (ETAT+) – a WHO recommended training package to support the poorest and most vulnerable children in rural and peri-urban areas.

[37] The training programme was complemented by the paediatric personnel employed to work in the hospital by the EBOVAC-Salone project to ensure appropriate clinical care for children enrolled in the Ebola vaccine trial. Improved clinical care has been a major benefit of hosting clinical trials in African communities but sustainability of the quality of care beyond the lifespan of the projects is usually a challenge. [38, 39]

Death was recorded in 7% of the children, the leading cause of which was severe malaria. Being an under-five child, wasted (i.e. WFH <-2SD), and having at least one danger sign were risk factors for death among the children. These factors resonate with findings of similar studies conducted in Africa [18, 22, 23] and underscore the need for improved nutritional support for children and better health-seeking behaviours of parents and caregivers of under-five children.

Contrary to the findings of a nation-wide survey on the impact of COVID-19 pandemic, which reported a slight reduction in the hospital utilisations but no significant changes in paediatric admissions across Sierra Leone [17], we observed a substantial reduction in the paediatric admissions during the COVID-19 pandemic, especially during the period shortly after the first national lockdown in Sierra Leone. The reason for this disparity is not known, but it might be connected with the design of the national survey which focused on a specific time point during the early phase of the pandemic. Although, children have been reported to carry a low risk of morbidity and mortality from COVID-19, reduction in the hospital admissions during COVID-19 pandemic led to planned health service re-configurations, with prioritisation of COVID-19 infection, and reduction of non-essential services, unplanned service disruption and changes in health-seeking behaviour. [17]

Our study had a few limitations. First, the clinical diagnoses were based almost entirely on clinical judgment with very limited laboratory support. Second, although it was documented that malaria was by far the most important cause of admission to the paediatric ward of Kambia hospital, clinical staff had not had enough experience, or had sufficient laboratory support, to allocate all hospital admissions with severe malaria to one of WHO recommended subgroups of this condition. Third, there were some missing data for anthropometric indices of the children. It is possible that these anthropometric measurements were taken by the ward staff, but were not recorded. Also, the proximity of the hospital to the homes of some children could not be estimated because their parents/caregivers did not provide specific home addresses, making it difficult to estimate the distance between their homes and the hospital. Nevertheless, the overall response rate of about 80% of children admitted to the ward during this study enabled us to demonstrate that the challenges of lack of systematic documentation of medical histories and poor record keeping of hospital admissions and outcomes can be overcome with a simple tool. Following successful implementation, the tool was adopted by the management of the hospital as the standard method of record keeping for the paediatric ward and has been adapted for other wards in the hospital.

## Conclusion

We developed and implemented a simple tool for documentations of paediatric admissions and outcomes at a rural hospital in a resource-limited setting. We found that severe malaria remains the leading cause of admission and mortality among the hospitalised children and advocate continuous use of the tool to improve paediatric care services delivery.

## Data Availability

Data for this study are accessible on Dryad: https://datadryad.org/stash/share/jJ04o5eI8WTbdEisPCjf_QypbL8xD8rGOKOUUZXF0eQ.

## Acknowledgements

We thank the staff and management of Kambia Government Hospital, Kambia District Health Management Team, the EBOVAC-Salone study team, the visiting paediatricians, expert trainers who supported the paediatric team, the COMAHS and LSHTM project management teams for logistical, administrative and management support. We thank all study participants and their families.

## Author contributions

MOA conceptualised the study and developed the tool. MOA, HHA and BK coordinated the data collection. YN, DK, ABJ, ES, and LO handled the data management. PA performed the statistical analysis. MOA wrote the first draft of the manuscript. BG supervised the study implementation, revised the tool and reviewed the early drafts of the manuscript. All authors reviewed the manuscript for substantial intellectual inputs and approved the final version of the manuscript for submission.

## List of figure and table legends

Supplementary file S1: Audit tool

